# Gendered time, financial & nutritional benefits from access to pay-as-you-go LPG for cooking in an informal settlement in Nairobi, Kenya

**DOI:** 10.1101/2022.06.02.22275930

**Authors:** Matthew Shupler, Jonathan Karl, Mark O’Keefe, Helen Hoka Osiolo, Tash Perros, Willah Nabukwangwa Simiyu, Arthur Gohole, Federico Lorenzetti, Elisa Puzzolo, James Mwitari, Daniel Pope, Emily Nix

## Abstract

**Introduction:** Few studies have examined gendered benefits of transitioning from polluting cooking fuels (e.g. charcoal, kerosene) to cleaner fuels (e.g. liquefied petroleum gas (LPG)). This study investigates pathways between adoption of pay-as-you-go (PAYG) LPG and women’s empowerment in Nairobi, Kenya.

**Methods:** Female (N=304) and male (N=44) primary cooks in an informal settlement in Nairobi were surveyed from December 2021-January 2022. The majority (84%; N=293) were customers of PayGo Energy, a company offering PAYG LPG. Other individuals (16%; N=55) cooking with full cylinder LPG or polluting fuels were randomly sampled from the community. The 45-minute telephonic survey examined how access to PAYG LPG affected the livelihoods of PayGo Energy’s customers.

**Results:** PayGo Energy customers were 50% more likely to cook exclusively with LPG (60%) than those using full cylinder LPG (40%). Due to reduced cooking times (average reduction: 42 min/day among previous polluting fuel users) from the adoption of PAYG LPG, the majority (58%; N=70) of female household heads took on additional employment compared with 36% (N=55) of females living in male-headed households. A greater proportion of married female household heads used their monetary savings from cooking with PAYG LPG for investment (41%) or savings (35%), compared with married women that were not household heads (3% and 21%, respectively). Increased dietary diversity and consumption of protein-rich foods (legumes, meat, fish) from cooking with PAYG LPG was reported by 15% of female household heads.

**Conclusion:** Female household heads were more likely than non-household heads to experience economic and nutritional gains when adopting PAYG LPG, illustrating how the agency of women influences their social co-benefits when undergoing clean energy transitions.

## INTRODUCTION

Over three billion individuals burn polluting fuels, such as wood, charcoal, and kerosene, in traditional stoves (e.g. open fires) for cooking, heating, and lighting. Household air pollution (HAP) generated from incomplete combustion of these fuels can lead to numerous adverse health conditions (Burroughs Pena et al., 2015; Kurmi et al., 2013; Smith et al., 2011; Thompson et al., 2011; Yu et al., 2018). Unsustainable harvesting of wood for cooking and charcoal production can result in deforestation (Anenberg et al., 2013).

In sub-Saharan Africa (SSA), use of polluting cooking fuels is most prevalent; approximately 85% of the population lacks access to clean fuels (Stoner et al., 2021). Across SSA, gathering wood for cooking and charcoal production accounted for nearly 50% of forest degradation in 2012 (Hosonuma et al., 2012). As a result of the negative health and climate impacts of reliance on polluting cooking fuels, several SSA countries are targeting rapid market expansion of liquefied petroleum gas (LPG) as a cleaner fuel for cooking (Bruce et al., 2017; Van Leeuwen et al., 2017). Although LPG is a fossil fuel, its use can have a neutral or cooling effect on climate when accounting for the reduction in emissions of CO_2_ and black carbon from replacement of biomass combustion in traditional stoves or open fires (Čukić et al., 2021; Kypridemos et al., 2020; Singh et al., 2017), and decreases in localized deforestation (Bailis et al., 2015). LPG also has a lower infrastructure requirement relative to electricity, making it more scalable in the short-term in SSA (Bruce et al., 2017; Chen et al., 2021).

### Pay-as-you-go liquefied petroleum gas

Pay-as-you-go (PAYG) LPG is an emerging model for the consumption of LPG whereby households are able to purchase LPG credits in small increments (via mobile banking) (Perros et al., 2021; Shupler et al., 2021c). Although there is a surcharge to the fuel costs to cover equipment installation and delivery fees, PAYG is a model that can provide resource-poor households in SSA with the payment flexibility needed to maintain clean cooking during periods of reduced household income (Shupler et al., 2021c). PAYG LPG companies track customers’ LPG consumption in real-time, which allows them to offer timely cylinder exchange through home deliveries before the customers run out of gas (Clean Cooking Alliance, 2019). This eliminates the need for customers to travel to LPG retail locations, which can be a significant LPG access barrier, particularly for women (Shupler et al., 2022b). PAYG can also improve LPG safety by minimizing the risk of illegal cylinder refilling due to increased transparency and efficiency of the LPG supply chain (O’Keefe and Marcigot, 2020).

PAYG LPG companies in East Africa have started operating in urban areas due to a greater population density allowing for easier distribution and logistics. Over half the urban population in SSA lives in informal settlements, which are overcrowded with insufficient housing, roads, water, electricity, and sanitation (Aboderin et al., 2017; Dianati et al., 2019). As families in informal settlements are resource poor and have limited access to clean energy sources (Parikh et al., 2012), they may benefit the most from lower upfront cost and flexible payment scheme for household energy. Women in particular may be socioeconomically empowered by PAYG LPG access as they face greater barriers to upward mobility than men in informal settlements (Prabhu, 2010) due to lower access to infrastructure services, and institutional and financial support (Parikh et al., 2015).

### Cooking is a gendered issue

Women are traditionally tasked with cooking in many LMICs (Johnson et al., 2019; Kumar and Mehta, 2016; Nguyen and Su, 2021) and may spend several hours per week collecting biomass fuels (Jagoe et al., 2020; Mazorra et al., 2020). This ‘time poverty’ is viewed as a key barrier to gender equality (Sustainable Development Goal 5) as it displaces time that women could otherwise use for employment or other productive activities (Petrokofsky et al., 2021). As informal settlements have been identified as particularly neglected in gender equality efforts (Musango et al., 2020), a transition to clean cooking fuels in these communities represents an important opportunity for empowering women (UN-Habitat, 2013).

Several studies have reported that cooking with LPG and electricity, rather than polluting fuels, can save women 30-60 minutes of cooking time per day (Krishnapriya et al., 2021; Maji et al., 2021; Simkovich et al., 2019). This reduction in cooking time from a clean energy transition may increase women’s productivity and, in turn, improve their well-being (O’Laughlin, 2007; Stevano, 2019); the link between access to LPG and improved mental health has been demonstrated in peri-urban sub-Saharan Africa (Shupler et al., 2022a) and rural India (Malakar and Day, 2020). However, how women use their newfound time savings (e.g. employment, professional development, domestic activities, leisure) is not widely researched (Petrokofsky et al., 2021).

### Contextualizing the gendered benefits of clean cooking access

When switching from polluting fuels to LPG, women and men can experience different benefits due to distinct socially constructed roles, responsibilities and needs (Musango, 2022). Moreover, the degree of benefits that women gain from adopting clean cooking fuels can be influenced by their level of autonomy and control over finances and other resources within their household (Kiptot et al., 2014; Yasmin and Grundmann, 2020). Thus, collecting data on gendered benefits of clean cooking access is critical for understanding how women’s agency influences their perceived benefits of clean energy access. Collecting empirical data on women’s empowerment can also improve future communication campaigns and implementation approaches and attract additional investment into the clean cooking sector by linking between universal access to clean energy (SDG 7), gender equality (SDG 5) and food security (SDG 2) (Shupler et al., 2021b).

Therefore, this study leverages data collected during the rapid expansion of PAYG LPG in East Africa to assess how household dynamics may influence women’s social and financial empowerment when undergoing the clean energy transition. Data on how PAYG LPG impacted livelihoods, including time and financial savings, stress levels and dietary patterns in an informal settlement in Nairobi, Kenya was disaggregated by sex of the household head to evaluate if women’s role in household decision-making impacted their perceived benefits.

## METHODS

### Study setting and population

This study took place in Nairobi, Kenya, where there are over 200 informal settlements that are home to over two million people (Beguy et al., 2015); informal settlements account for over 60% of Nairobi’s population yet only occupy about 5% of the city’s land (Ahmed et al., 2015; Mberu et al., 2016). The study is set within one of the largest clusters of informal settlements in Nairobi, Mukuru (home to over 150,000 families), which is located in Embakasi South and Makadara sub-counties of Nairobi on 650 acres of privately and publicly owned land in an industrial area (Kim et al., 2019). This study is based in four neighboring informal settlements within Mukuru: kwa Reuben, kwa Njenga, Viwandani and Juakali.

Mukuru informal settlement is the service area for PayGo Energy, a company offering PAYG LPG services in Nairobi since 2016. PayGo Energy provides a new double-burner stove, gas cylinder, smart meter, fire safety equipment and home delivery of cylinder refills to its customers. While PayGo Energy has residential and small commercial business customers, only PayGo Energy’s residential customers were included in this study to examine household-level cooking patterns and behaviors. Individuals who registered with PayGo Energy by August 2021 formed the sampling frame of PAYG LPG users for this study.

### Data collection and analysis

At the start of the study PayGo Energy’s customers’ contact information was securely shared with the research team via Google cloud. A total of 457 of PayGo Energy’s customers in the database were called by the field team trained by Busara Center for Behavioral Economics, a laboratory for behavioral and experimental economics based in Nairobi, Kenya (Haushofer et al., 2014). To evaluate potential socioeconomic benefits of access to PAYG LPG, a comparison group consisting of a random sample of 90 individuals not cooking with PAYG LPG were also included in the sampling frame. These participants were selected from a participant pool of over 5,000 residents of Nairobi’s informal settlements by Busara Center for Behavioral Economics. Among both the sample of PayGo Energy customers and the comparison group, 36% (n=198) of participants that were not the primary cook of the household were excluded from analysis to minimize potential biases associated with the community members’ cooking experiences and perceptions of the benefits of access to PAYG LPG.

Prior to data collection, researchers from Busara Center for Behavioral Economics developed, translated and piloted survey questions to ensure they were culturally appropriate. After finalizing the surveys, the recruited field team was trained on the aims of the study. Data was collected on smartphones using the ‘Collect’ smartphone application developed by Open Data Kit (ODK). Data was securely transferred from the smartphones to the ODK Cloud for storage. The ODK cloud uses Amazon Web Services (AWS) for their infrastructure, which has end-to-end encryption.

Prior to enrollment, individuals were informed about the aims of the study, that their participation was voluntary, that they could choose to stop the survey at any time and that their data would be kept confidential. All participants completed a 45-minute telephonic, cross-sectional survey from December 2021-January 2022. The survey documented potential social and economic benefits of adopting PAYG LPG, including time savings, financial gains, food security (survey question: ‘over the past three months, how often have you or your household had to skip a meal because you didn’t have food?’), dietary diversity (survey question: ‘How did pay-as-you-go LPG affect the diversity of food you cook?’), consuming a balanced diet (survey question: ‘How often per week do you eat a balanced diet (one meal per day containing carbohydrates (such as ugali or rice), proteins (such as meat or beans) and vegetables)?’), consuming a healthy diet (survey question: ‘Is your diet now more or less healthy than before using pay-as-you-go LPG?) and impacts on stress (survey question: ‘Has using pay-as-you-go LPG reduced the amount of stress in your daily life?’). Descriptive statistics of associations between these socioeconomic benefits and the sex of the household head are presented.

All data analyses were conducted in R version 4.1.1 (R Core Team, 2017). Ethical approval was obtained from the National Commission for Science, Technology & Innovation (NACOSTI) in Nairobi, Kenya. Informed verbal consent was obtained from all participants prior to conducting the study.

## RESULTS

A total of 348 primary cooks were included in the final analytic sample, including 293 PayGo Energy customers and 55 primary cooks not using PAYG LPG (n=40 used full cylinder LPG). The vast majority (85%; N=249) of PayGo Energy customers were female, with half (51%; N=126) being the head of household and half living in a household run by their male partner (49%; N=123). Male primary cooks that were PayGo Energy customers were almost always (98%; N=44) also the household head.

Almost all (98%; N=120) of female PayGo Energy customers living in male-headed households were married, compared with just over half (53%; N=67) of female PayGo Energy customers that were the household head (Table 1).

**Table 1.** Socioeconomic characteristics of PAYG LPG customers by sex of the household head and primary cook (N=293)

|  | **Overall**<br>**(N=293)** | **Female household**<br>**head & female**<br>**primary cook**<br>**(N=126)** | **Male household**<br>**head & female**<br>**primary cook**<br>**(N=123)** | **Male household**<br>**head & male**<br>**primary cook**<br>**(N=44)** |
| --- | --- | --- | --- | --- |
| Age (Mean (SD)) | 38 (10) | 38 (10) | 40 (9) | 35 (11) |
| Marital status |  |  |  |  |
| Single | 98 (33%) | 67 (53%) | 3 (2%) | 28 (64%) |
| Married | 170 (58%) | 35 (28%) | 120 (98%) | 15 (34%) |
| Divorced or separated | 13 (5%) | 13 (10%) | 0 | 0 |
| Widowed | 12 (4%) | 11 (9%) | 0 | 1 (2%) |
| In charge of cooking fuel decisions |  |  |  |  |
| Participant | 233 (82%) | 120 (98%) | 80 (66%) | 33 (80%) |
| Participant & spouse | 44 (16%) | 2 (2%) | 35 (29%) | 7 (17%) |
| Spouse | 7 (2%) | 0 | 6 (5%) | 1 (3%) |
| Primary source of income |  |  |  |  |
| Self-employed | 146 (51%) | 66 (54%) | 63 (52%) | 17 (40%) |
| Formal employment | 62 (22%) | 24 (20%) | 20 (16%) | 18 (43%) |
| Informal work | 73 (25%) | 31 (25%) | 35 (29%) | 7 (17%) |
| None | 6 (2%) | 2 (1%) | 4 (3%) | 0 |
| Monthly income |  |  |  |  |
| ≤10,000 KES<sup>1</sup> | 62 (22 %) | 37 (30%) | 19 (16%) | 6 (14%) |
| 10,001-20,000 KES | 115 (40%) | 52 (42%) | 47 (38%) | 16 (38%) |
| >20,000 KES | 110 (38%) | 34 (28%) | 56 (46%) | 20 (48%) |
| Income frequency |  |  |  |  |
| Daily | 130 (45%) | 60 (49%) | 60 (49%) | 10 (24%) |
| Weekly | 20 (7%) | 12 (10%) | 5 (4%) | 3 (7%) |
| Monthly | 86 (30%) | 29 (24%) | 32 (26%) | 25 (60%) |
| Irregular/unpredictable | 51 (18%) | 22 (18%) | 25 (21%) | 4 (9%) |
| Urgent financial expenses (multiple responses allowed) |  |  |  |  |
| Rent | 267 (91%) | 114 (91%) | 113 (92%) | 39 (89%) |
| Education/school fees | 205 (70%) | 81 (64%) | 98 (80%) | 25 (57%) |
| Utility bills | 88 (30%) | 36 (29%) | 34 (28%) | 17 (39%) |
| Cooking fuel | 57 (19%) | 29 (23%) | 16 (13%) | 12 (27%) |
| Medical expenses | 54 (18%) | 28 (22%) | 20 (16%) | 6 (14%) |
| Savings | 32 (11%) | 16 (13%) | 9 (7%) | 7 (16%) |
| Business expenditures | 25 (9%) | 13 (10%) | 8 (7%) | 4 (9%) |
| Food insecurity during last 3 months | 140 (48%) | 67 (54%) | 56 (46%) | 16 (36%) |
| Family size |  |  |  |  |
| 1-2 | 29 (13%) | 14 (15%) | 5 (5%) | 9 (25%) |
| 3-4 | 119 (53%) | 50 (53%) | 46 (50%) | 23 (64%) |
| 5+ | 75 (34%) | 30 (32%) | 41 (45%) | 4 (11%) |
| Other cooking fuels used at time of survey (multiple options allowed) |  |  |  |  |
| None (LPG only) | 175 (60%) | 77 (61%) | 61 (50%) | 37 (84%) |
| Charcoal | 77 (26%) | 35 (24%) | 37 (29%) | 5 (11%) |
| Kerosene | 42 (14%) | 16 (9%) | 24 (19%) | 2 (4%) |
| Electricity | 12 (4%) | 3 (2%) | 7 (6%) | 2 (4%) |
| Ethanol | 3 (1%) | 2 (2%) | 1 (1%) | 0 |
| Wood | 3 (1%) | 2 (2%) | 1 (1%) | 0 |
| Previous primary cooking fuel(s) |  |  |  |  |
| LPG (full cylinder) | 215 (73%) | 88 (70%) | 92 (75%) | 35 (80%) |
| Kerosene | 38 (13%) | 16 (13%) | 15 (12%) | 7 (16%) |
| Charcoal & kerosene | 30 (10%) | 17 (13%) | 12 (10%) | 1 (2%) |
| Charcoal | 9 (3%) | 4 (3%) | 4 (3%) | 1 (2%) |
| Amount spent (KES<sup>1</sup>) weekly on all cooking fuels (median (IQR)) | 300 [200,500] | 300 [200,488] | 350 [225,550] | 200 [100, 400] |

1. KES= Kenyan Shilling

### Comparing socioeconomic characteristics of PayGo Energy customers by sex of the household head

Almost all (98%; N=120) female customers of PayGo Energy that were the household head reported overseeing cooking fuel-related decisions, compared with two-thirds (66%; N=80) of women that were not the household head (Table 1). However, in households headed by males, 95% of women reported playing a role in cooking-related decision-making (Table 2).

**Table 2.** Socioeconomic characteristics between customers and noncustomers of PayGo Energy by sex of the household head (female primary cooks only)

|  | PayGo Energy customers |  | Non PayGo Energy customers |  |
| --- | --- | --- | --- | --- |
|  | Female household head (N=126) | Male household held (N=123) | Female household held (N=27) | Male household head (N=28) |
| Age (Mean (SD)) | 38 (10) | 40 (9) | 40 (10) | 38 (10) |
| Marital status |  |  |  |  |
| Single | 67 (53%) | 3 (2%) | 8 (31%) | 2 (7%) |
| Married | 35 (28%) | 120 (98%) | 5 (19%) | 25 (93%) |
| Divorced or separated | 13 (10%) | 0 | 6 (23%) | 0 |
| Widowed | 11 (9%) | 0 | 7 (27%) | 0 |
| Primary source of income |  |  |  |  |
| Self-employed | 66 (54%) | 63 (52%) | 14 (52%) | 12 (44%) |
| Formal employment | 24 (20%) | 20 (16%) | 2 (7%) | 5 (19%) |
| Informal work | 31 (25%) | 35 (29%) | 11 (41%) | 9 (33%) |
| None | 2 (1%) | 4 (3%) | 0 | 1 (4%) |
| Monthly income |  |  |  |  |
| <=10,000 KES<sup>1</sup> | 37 (30%) | 19 (16%) | 10 (37%) | 4 (15%) |
| 10,001-20,000 KES | 52 (42%) | 47 (38%) | 14 (52%) | 14 (52%) |
| >20,000 KES | 34 (28%) | 56 (46%) | 3 (11%) | 9 (32%) |
| Income frequency |  |  |  |  |
| Daily | 60 (49%) | 60 (49%) | 18 (67%) | 12 (43%) |
| Weekly | 12 (10%) | 5 (4%) | 1 (4%) | 0 |
| Monthly | 29 (24%) | 32 (26%) | 3 (11%) | 6 (21%) |
| Irregular/unpredictable | 22 (18%) | 25 (21%) | 5 (18%) | 9 (32%) |
| Food insecurity during last 3 months | 67 (54%) | 56 (46%) | 17 (63%) | 7 (25%) |
| Family size |  |  |  |  |
| 1-2 | 14 (15%) | 5 (5%) | 2 (11%) | 1 (6%) |
| 3-4 | 50 (53%) | 46 (50%) | 8 (44%) | 10 (63%) |
| 5+ | 30 (32%) | 41 (45%) | 8 (44%) | 5 (31%) |
| Other cooking fuels used at time of survey (multiple options allowed) |  |  |  |  |
| None (LPG only) | 77 (61%) | 61 (50%) | 9 (33%) | 7 (25%) |
| Charcoal | 35 (24%) | 37 (29%) | 11 (41%) | 8 (29%) |
| Kerosene | 16 (9%) | 24 (19%) | 9 (33%) | 8 (29%) |
| Electricity | 3 (2%) | 7 (6%) | 1 | 0 |
| Ethanol | 2 (2%) | 1 (1%) | 1 (3%) | 1 (3%) |
| Wood | 2 (2%) | 1 (1%) | 0 | 2 (7%) |
| In charge of cooking fuel decisions |  |  |  |  |
| Participant | 120 (98%) | 80 (66%) | 24 (89%) | 5 (19%) |
| Participant & spouse | 2 (2%) | 35 (29%) | 3 (11%) | 18 (67%) |
| Spouse | 0 | 6 (5%) | 0 | 4 (15%) |
| Amount spent (KES) weekly on all cooking fuels (Median [inner quartile range]) | 300 [200,500] | 300 [200,488] | 350 [200,525] | 312 [133,425] |

1. KES= Kenyan Shilling

Male PayGo Energy customers were twice as likely (43%) to be formally employed as female PayGo Energy customers (20%). Due to differences in employment type, male PayGo Energy customers were half as likely (24%) as female primary PayGo Energy customers (49%) to report receiving daily wages.

Female heads of household reported lower average household income (30% in lowest income bracket (<10,000 Kenyan Shillings (KES)) and 28% in highest income bracket (>20,000 KES)) than females living in male-headed households (16% in lowest income bracket; 46% in highest income bracket) and male primary cooks (14% in lowest income bracket; 48% in highest income bracket) (Table 1). Similarly, female headed households were more likely to be food insecure during the three months prior to the survey (54%) than male headed households (46%). Households with male primary cooks reported even lower levels of food insecurity (36%).

Almost two-thirds (N=175; 60%) of PayGo Energy’s customers reported cooking exclusively with PAYG LPG. Households with 1-2 individuals were more likely (76%; N=22) to cook exclusively with PAYG LPG than larger families with 3-4 members (58%; N=70) or 5-6 inhabitants (56%; N=42). Female primary cooks that were the household head were more likely (61%) to cook exclusively with PAYG LPG than female primary cooks that were not the head of the household (50%). Male cooks had the highest rate (84%) of exclusive PAYG LPG use. Participants cooking with other fuels alongside PAYG LPG mainly cooked with charcoal (26%; N=77) or kerosene (14%; N=42) (Table 1). Three-quarters (N=387; 73%) of PayGo Energy’s customers reported previously cooking with full cylinder LPG before switching to PAYG LPG (Table 1).

### Comparing socioeconomic characteristics between PayGo Energy customers and the general community

Among all female primary cooks in the informal settlement, half (N=153) were the household head (Table 2); the proportion of women with control over household decisions did not vary between customers of PayGo Energy and non-customers (Table 2). However, female household heads cooking with PAYG LPG were more highly educated (29% primary level education or less) than those not using PAYG LPG (59% primary level education or less) (Figure 1). Furthermore, the proportion of female household heads working in the formal sector was three times as high among PAYG LPG users (20%) as non-users (7%) (Table 2). There was only a 3% difference in the prevalence of formal work status among those cooking with PAYG LPG (24%) versus those not (19%) among women living in male headed households.

**Figure 1.**
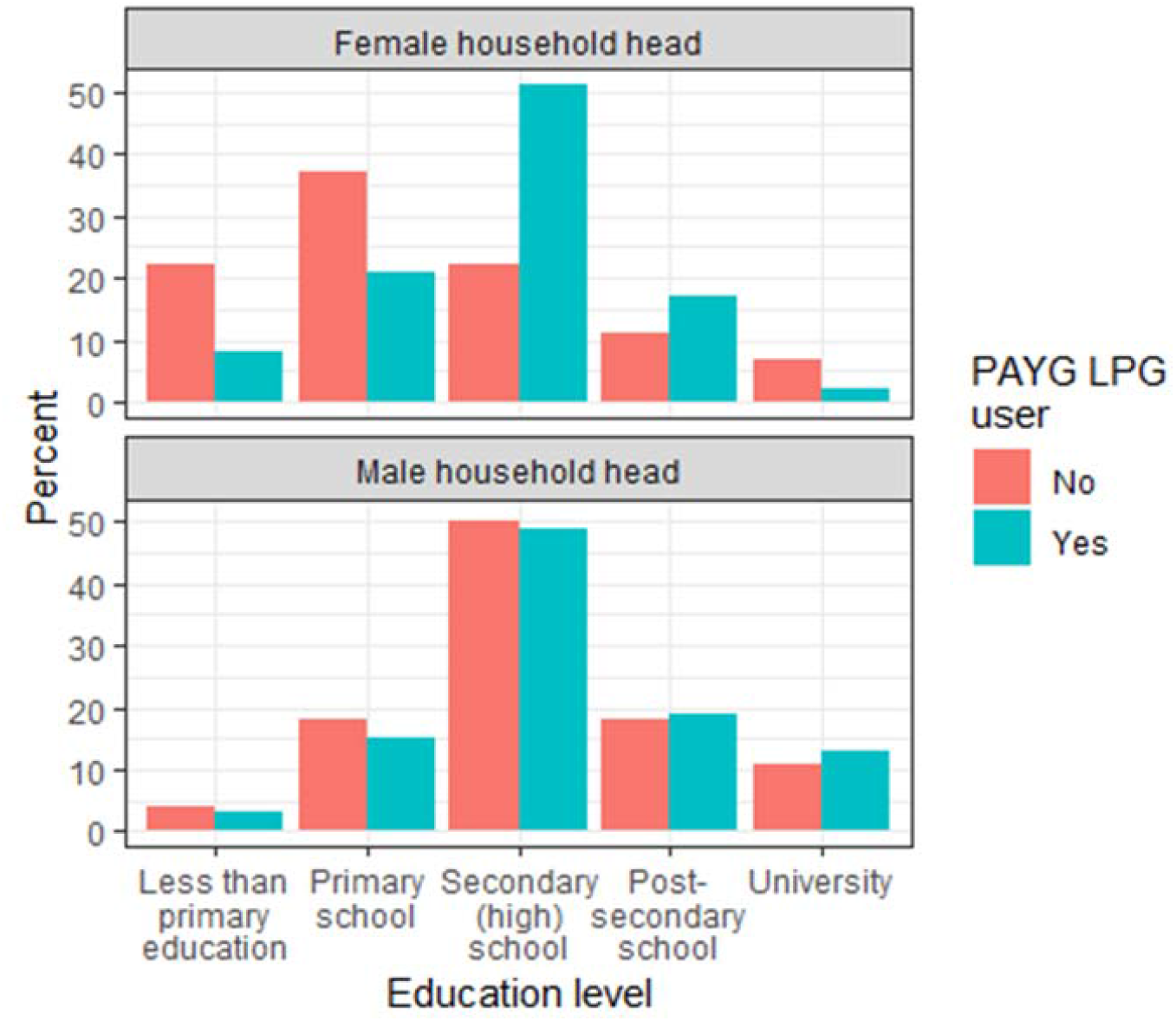
Education level among PAYG LPG users and the general community by sex of the household head

Among PAYG LPG customers, rates of food insecurity during the three months prior to the survey were 10% higher among households with female primary cooks. Among non-PAYG LPG customers, the prevalence of food insecurity was 2.5 times higher among households with female primary cooks (63%) as male primary cooks (25%).

Over half (54%) of female household heads using PAYG LPG were single compared with one-third (31%) of female household heads not cooking with PAYG LPG (Table 2). Female household heads that were not using PAYG LPG reported lower average household income (11% in the highest income bracket) and a higher prevalence of food insecurity (63%) than female heads using PAYG LPG (28% in highest income bracket; 54% food insecure).

Exclusive use of LPG was 50% higher among PayGo Energy’s customers (60%) than among those using full cylinder LPG in the general community (40%), irrespective of the sex of the household head. Additionally, female authority in cooking-related decisions was over three times higher among women cooking with PAYG LPG (66%) compared with those not (19%).

The proportion of female household heads using PAYG LPG who indicated that paying for cooking fuels was an urgent household expense (23%) was 14% lower than the proportion of female heads not using PAYG LPG (37%). However, a similar proportion of females cooking (13%) and not cooking (14%) with PAYG LPG in male headed households reported that the cost of cooking fuels was a major household expense.

### Time saved when cooking with pay-as-you-go liquefied petroleum gas

Nearly all (99%; N=67) PayGo Energy customers that switched from primary use of polluting fuels to PAYG LPG reported time savings. Even among households switching from full cylinder LPG to PAYG LPG, four fifths (82%; N=142) reported savings in time. The proportion of women reporting time savings did not differ by the sex of household head.

Self-reported reductions in cooking time among PayGo Energy’s customers that previously cooking with polluting fuels (42 min/day saved (ranging from 25 min/day to 67 min/day)) were greater than those who previously used full cylinder LPG (20 min/day saved (ranging from 26 min/day to 46 min/day)) (Figure 2).

**Figure 2.**
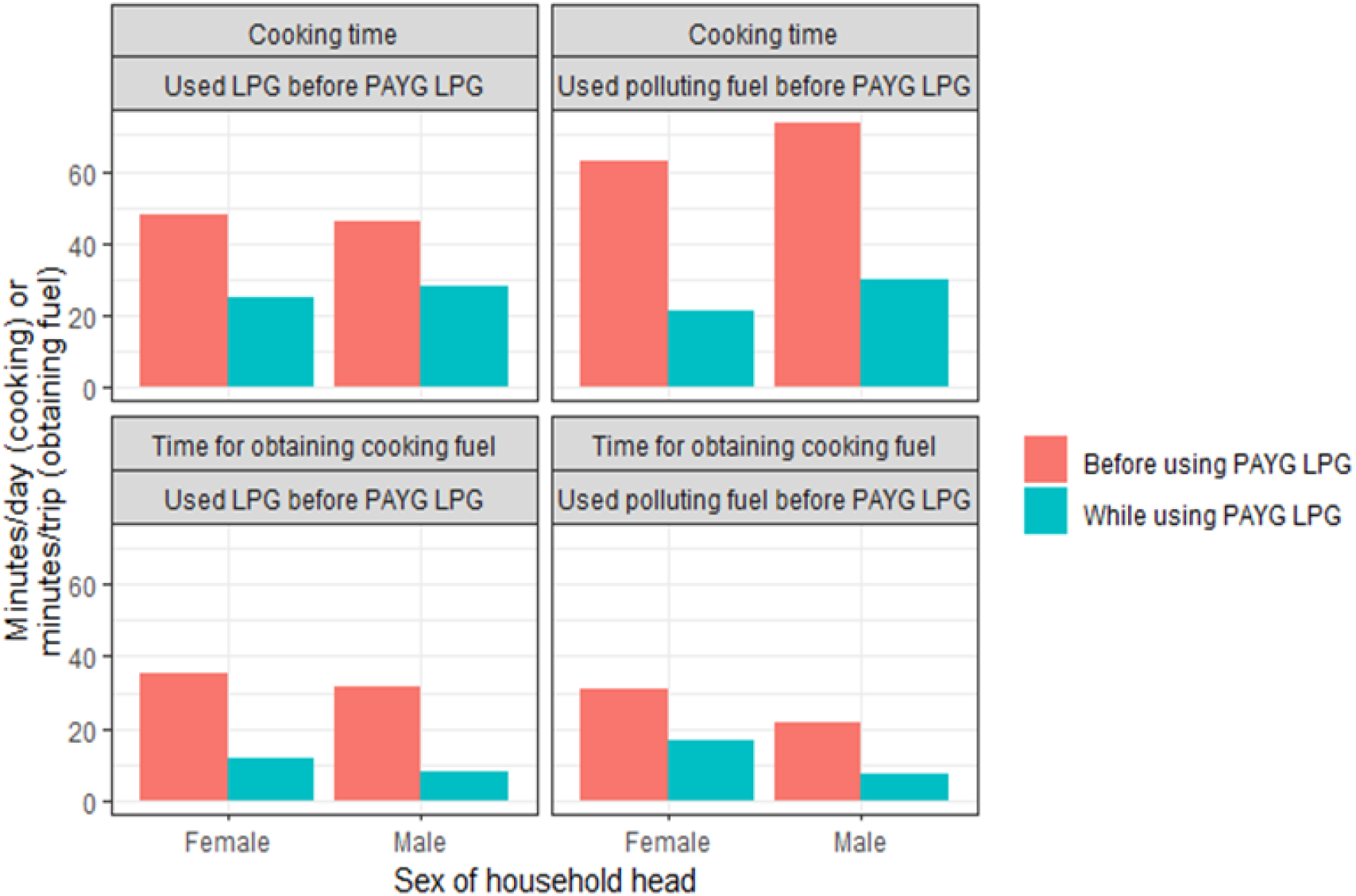
Time savings from using PAYG LPG by sex of the household head and previous primary cooking fuel

Women primarily cooking with full cylinder LPG prior to using PAYG LPG reported a 5-minute longer average travel time (33 minutes/trip) to obtain their cooking fuel than those only cooking with polluting fuels (28 minutes/trip). Thus, significant time savings from fuel collection were reported by participants regardless of their previous cooking fuel. Female primary cooks previously cooking with LPG also benefited from a 23-minute/trip average reduction (33 minutes to 10 minutes) in average time required to obtain their cooking fuels when using PAYG LPG; this reduction was longer than the 15-minute/trip average decrease (28 minutes to 13 minutes) in travel time reported by women cooking with polluting fuels prior to using PAYG LPG.

Among women previously cooking exclusively with polluting fuels, three-quarters (74%) of female household heads reported using their saved time to take on new employment (Figure 3). For women cooks living with male household heads, less than half (42%) used this time to take on new employment. Likewise, among women cooking with full cylinder LPG prior to registering with PayGo Energy, over half (52%) of household heads used their newfound time to work another job, compared with one-third (33%) of women that were not in charge of household decisions.

**Figure 3.**
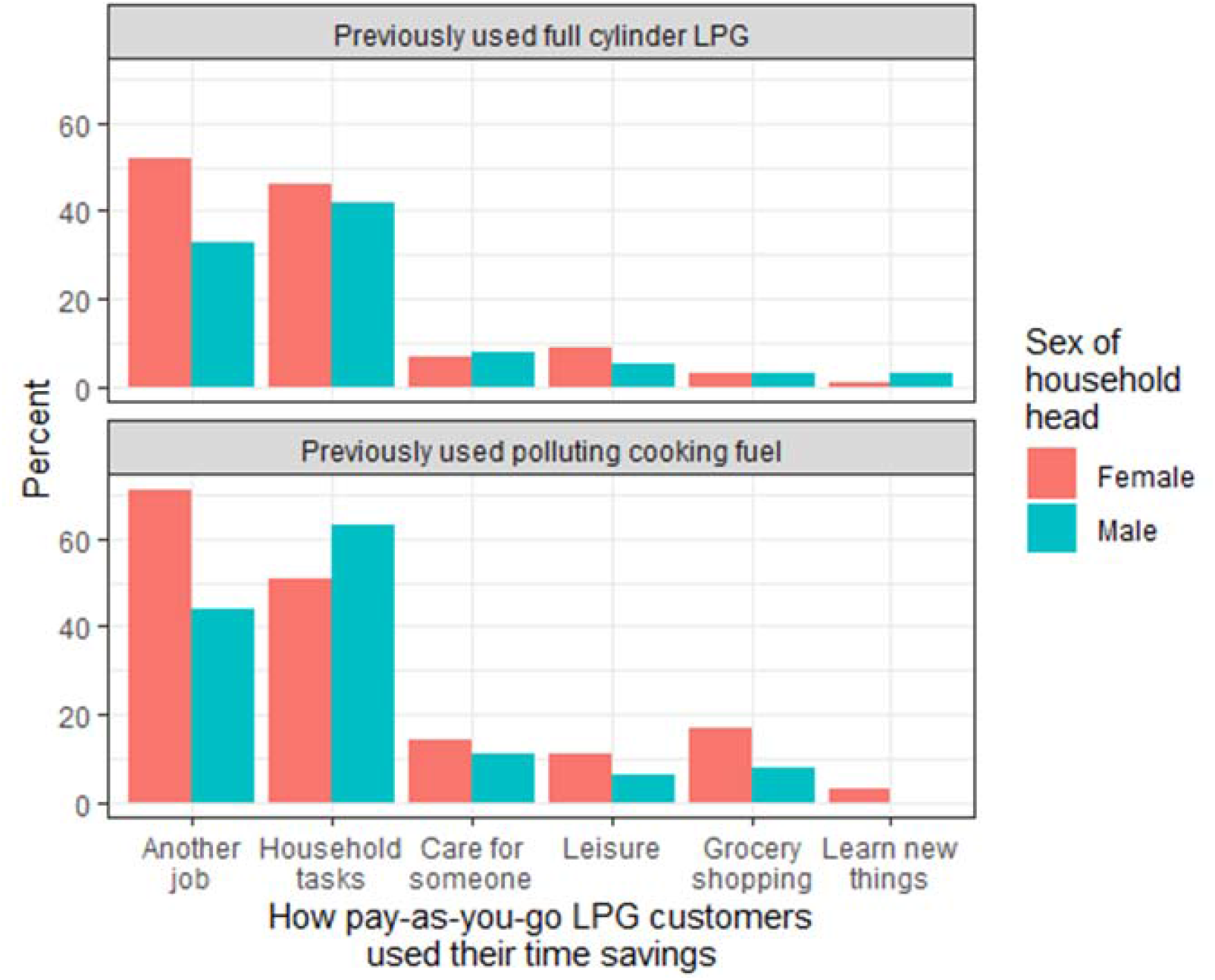
How pay-as-you-go LPG female customers used their time savings by sex of the household head and previous primary cooking fuel

Women living in male-run households that previously cooked with polluting fuels were more likely to reapportion their time from cooking to other household tasks; 63% reported taking on additional household tasks compared with 51% of female household heads. Only 7% (N=22) of women used their newfound time for leisure and hardly any (3%, N=8) used their time for educational purposes.

### Financial savings

In addition to time savings, over half (58%; N=145) of female primary cooks reported that switching to PAYG LPG for cooking saved them money. A higher proportion of women previously cooking with polluting fuels (82%; N=56) reported saving money compared with women formerly cooking with full cylinder LPG (51%, N=89).

A significant proportion of married women that were the head of household used their monetary savings from cooking with PAYG LPG for investment (41%) or savings (35%). Unmarried female household heads used their monetary gains to a lesser extent for investment (9%) or savings (23%). A lower proportion of married women living in male headed households used their financial savings for investment (3%) or savings (21%) (Figure 4). Among married women, those that were not the head of household had a 27% greater probability of spending their earnings from use of PAYG LPG (80%) than female household heads (53%).

**Figure 4.**
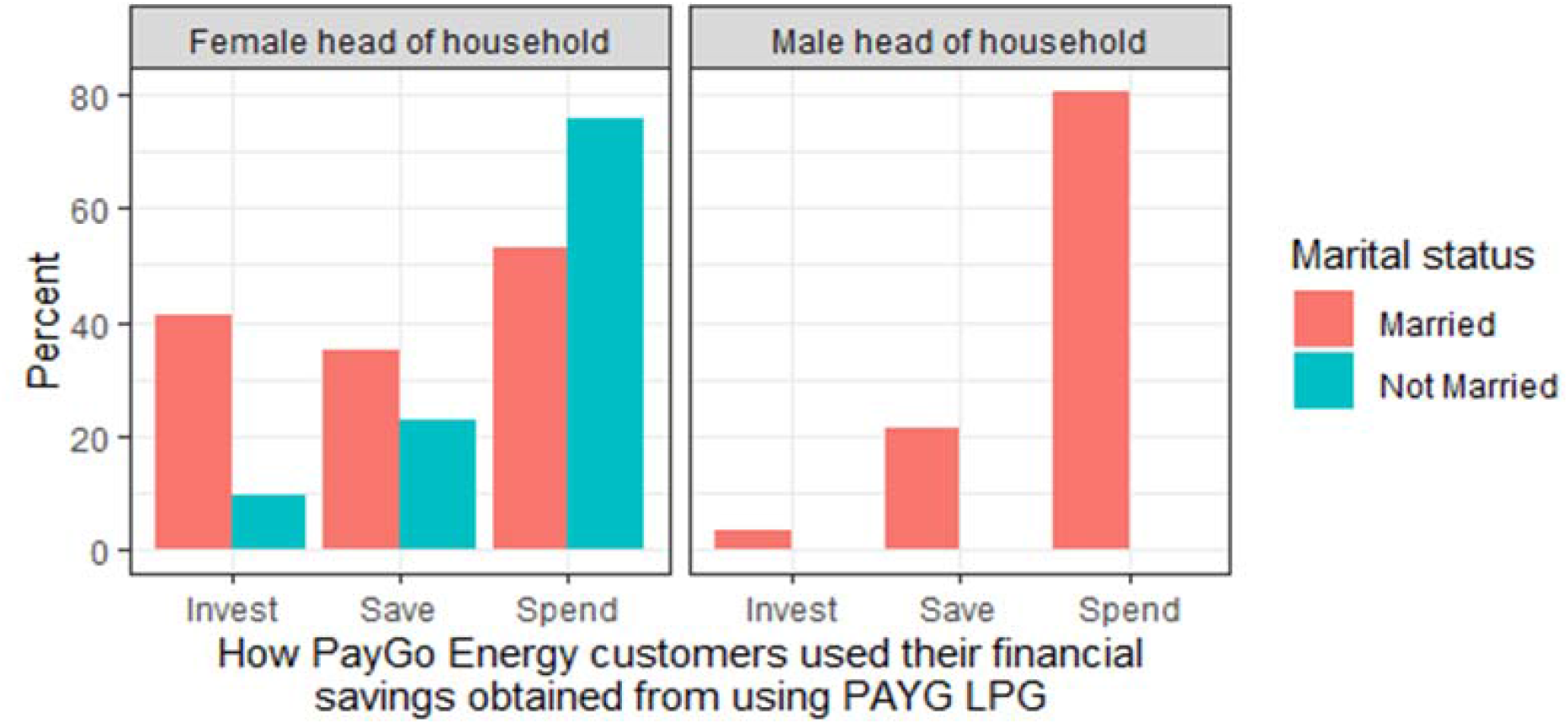
How pay-as-you-go LPG customers used their financial savings, stratified by sex of the household head and marital status

### Dietary changes when using pay-as-you-go LPG

The prevalence of female household heads reporting that their diet was healthier (16%; N=19) or more diverse (15%; N=18) as a result of cooking with PAYG LPG was nearly double that of women living in male-run households (8% (N=9) and 8% (N=9), respectively) (Figure 5). Female household heads consistently reported consuming more carbohydrates, legumes, fish and meat as a result of using PAYG LPG compared with women living in households headed by males. The largest discrepancies between the sex of household heads in changes to the type of food consumption was among protein sources: female household heads were three times as likely to report preparing more fish (12%; N=12) and more meat (15%; N=18) as a result of cooking with PAYG LPG than women living in male-run households (fish: 5%; N=5 meat: 3%; N=3) (Figure 5).

**Figure 5.**
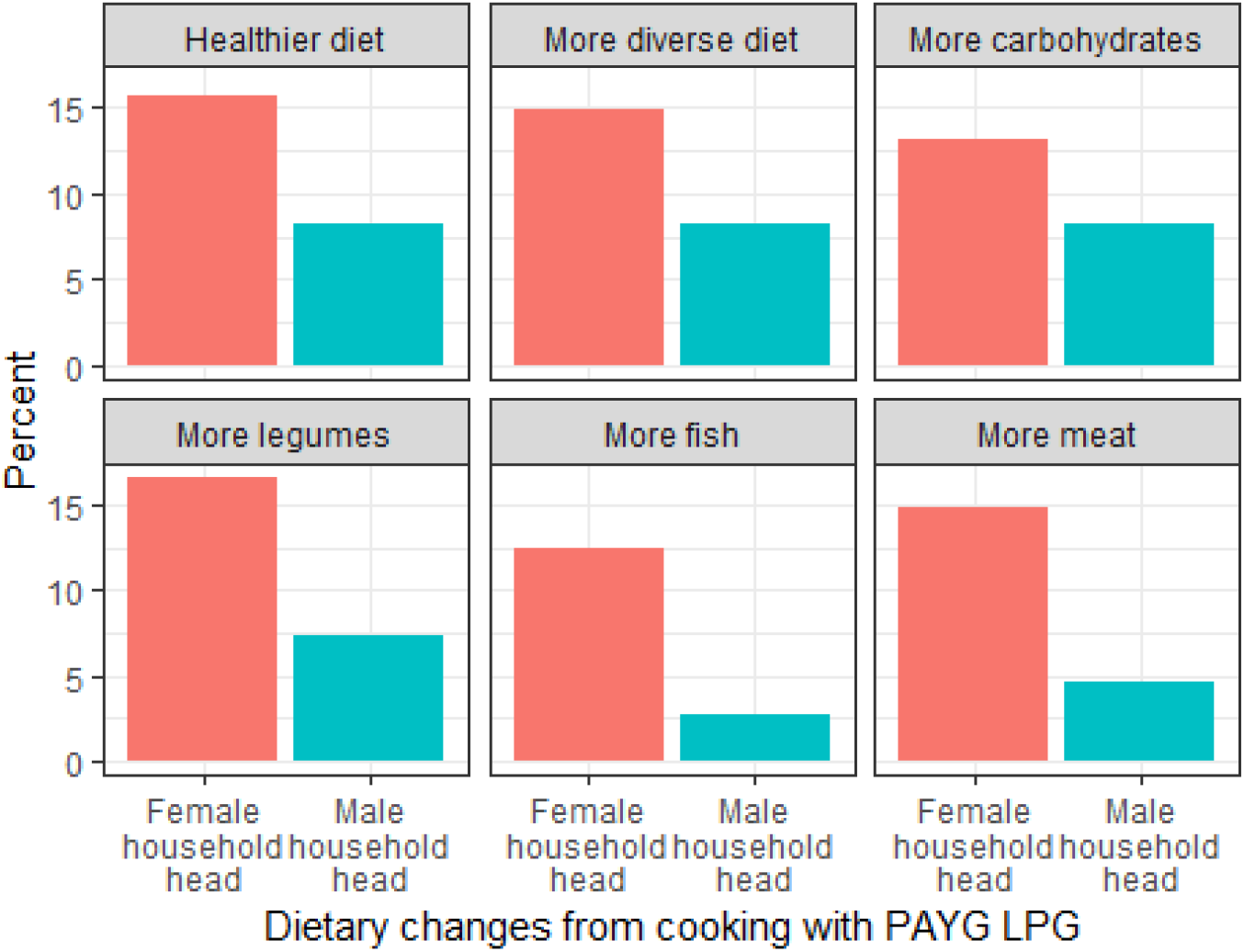
Proportion of women reporting dietary changes stratified by sex of the household head

### Stress relief and fulfilled aspirations from using pay-as-you-go liquefied petroleum gas

Food insecurity and household expenses were a greater source of stress for female household heads compared to male household heads (Figure 6). Among female household heads cooking with PAYG LPG,, the prevalence of all causes of stress included in the survey were significantly (5-15%) lower compared to those not cooking with PAYG LPG. Contrastingly, among women that were not the head of household, sources of stress related to finances and food security were higher for those cooking with PAYG LPG compared to those not cooking with PAYG LPG (Figure 6).

**Figure 6.**
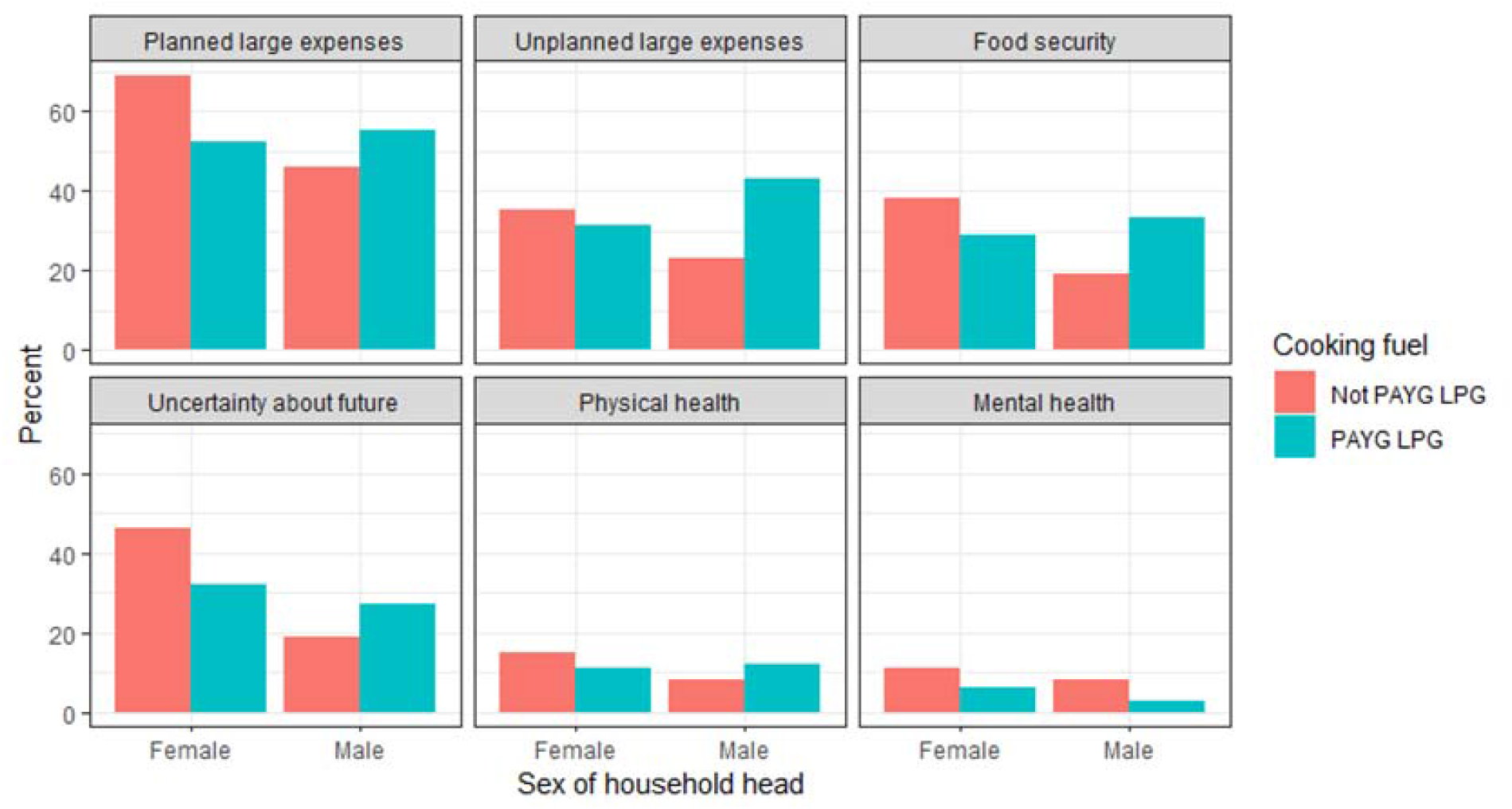
Sources of stress among primary cooks by sex of the household head and whether they cook with pay-as-you-go LPG

Seven out of ten women (N=169) in the study reported that their life was ‘stressful’; rates of stress were approximately 5-10% lower among females that were heads of households versus those that were not (Table 3). Nearly half (45%) of PayGo Energy customers believed that cooking with PAYG LPG had allowed them to fulfill other aspirations which they could not pursue previously (Table 3). Among women cooking with PAYG LPG that previously did not cook with full cylinder LPG, 60% stated that they were able to fulfill previous aspirations; over one-third (38%) of women that had previously cooked with full cylinder LPG reported having previous aspirations fulfilled because of cooking with PAYG LPG (Table 3).

**Table 3.** Impact of using PAYG LPG on stress levels and aspirations of women by sex of household head and previous cooking fuel

|  | All female<br>PAYG LPG<br>users<br>(N=243) | Previously cooked with full<br>cylinder LPG |  | Previously did not cook with<br>full cylinder LPG |  |
| --- | --- | --- | --- | --- | --- |
|  |  | Female<br>household<br>head<br>(N=82) | Male<br>household<br>head<br>(N=80) | Female<br>household<br>head<br>(N=38) | Male<br>household<br>head<br>(N=27) |
| Life is stressful |  |  |  |  |  |
| Yes | 169 (70%) | 50 (61%) | 56 (70%) | 26 (73%) | 21 (78%) |
| Stress reduced by using PAYG<br>LPG | 143 (64%) | 50 (62%) | 56 (71%) | 19 (54%) | 19 (67%) |
| PAYG LPG allowed previously<br>unpursued aspirations to be fulfilled | 99 (45%) | 31 (39%) | 29 (37%) | 20 (57%) | 19 (73%) |

### Other perceived benefits of pay-as-you-go LPG

While the ability to pay for gas in smaller incremental payments was the main reason for recommending PAYG LPG to others (83%), the convenience of PAYG LPG in terms of the provision of a double burner stove (in contrast to the single burner stove typically used by individuals cooking with full cylinder LPG in the community) and subsequent home delivery of cylinder refills was also highly valued (73%) in terms of recommending the technology (83%) (Table 4). Improved health was less frequently mentioned (29%) as a benefit that PayGo Energy’s customers would recommend PAYG LPG to other community members.

**Table 4.** Features of PAYG LPG favored by PayGo Energy’s customers

|  | **Overall**<br>**(N=294)** | **Female primary cook** |  | **Male**<br>**primary**<br>**cook**<br>**(N=44)** |
| --- | --- | --- | --- | --- |
|  |  | **Female head**<br>**of household**<br>**(N=126)** | **Male head of**<br>**household**<br>**(N=123)** |  |
| Reasons for recommending PAYG LPG to others (multiple responses allowed) |  |  |  |  |
| Small payments | 243 (83%) | 103 (82%) | 105 (85%) | 35 (80%) |
| Convenience | 218 (73%) | 90 (71%) | 99 (80%) | 29 (66%) |
| Overall cost | 127 (44%) | 60 (48%) | 49 (40%) | 18 (41%) |
| Security of having cooking fuel | 89 (31%) | 34 (27%) | 40 (32%) | 15 (34%) |
| Better health | 85 (29%) | 41 (33%) | 29 (24%) | 15 (34%) |
| Started using PayGo Energy because someone else in community was using it | 126 (45%) | 62 (51%) | 46 (38%) | 18 (44%) |
| PAYG LPG users enjoy more social recognition in the community | 182 (62%) | 87 (71%) | 65 (54%) | 30 (73%) |
| Prefer to purchase LPG in bulk or small amounts |  |  |  |  |
| Bulk | 106 (36%) | 50 (39%) | 40 (33%) | 16 (36%) |
| Small amounts | 180 (61%) | 74 (59%) | 78 (63%) | 28 (64%) |
| No preference | 7 (3%) | 2 (2%) | 5 (4%) | 0 |

Although there were not substantial differences by sex (cook and household head) in customers’ most recommended features of PAYG LPG, a greater proportion of female household heads (73%) and male primary cooks (71%) believed that PAYG LPG users enjoyed greater social recognition in the community compared with women in male headed households (54%) (Table 4). Moreover, female primary cooks that were the household head were more likely (51%) to start cooking with PAYG LPG because others in the community were using the service than women that were not in charge of household decisions (38%).

Almost two-thirds (61%) of participants preferred purchasing LPG in smaller increments compared to buying in bulk (full cylinder) (36%); there were no major differences in preferences between sexes or sex of the household head (Table 4).

## DISCUSSION

This study explores the gendered dimensions of PAYG LPG adoption in an informal urban settlement in Nairobi, Kenya. Building on other research that has linked access to clean cooking fuels to women’s empowerment (Maji et al., 2021; Nguyen and Su, 2021; Yasmin and Grundmann, 2019), we find that women cooking with PAYG LPG reported improvements in multiple aspects of their livelihoods: lower stress levels (Figure 6), greater consumption of protein-rich foods and more diverse diets (Figure 5), and substantial financial (Figure 4) and time savings (Figure 2) enabling them to take on additional employment (Figure 3).

Social and economic gains from adopting PAYG LPG for cooking were reported by a large proportion of female primary cooks in the study community. However, the use of PAYG LPG, and therefore the extent of the perceived benefits, varied based on the female cooks’ status within the household. Female primary cooks that were also the household head were more likely to cook exclusively with PAYG LPG (61%) than female primary cooks who were not (50%). Additionally, among male headed households, women in charge of cooking-related decisions were more than three times more likely to cook exclusively with PAYG LPG (66%) than women who lacked authority over decisions related to cooking fuels (19%). A much higher proportion of female household heads (74%) who previously cooked with polluting fuels used their time savings to start new employment, compared with their counterparts who lived in households headed by males (42%) (Figure 3). This association also held among women switching from full cylinder LPG to PAYG LPG: a larger percentage of household heads (52%) relative to women not in charge of household decisions (33%) reported starting another job with their saved time. These findings underscore the underlying gendered dynamics within informal urban settlements in SSA.

### Socioeconomic differences between PayGo Energy customers and the rest of the community

When comparing PAYG LPG customers to the general community, socioeconomic characteristics differed only among female headed households; female household heads cooking with PAYG LPG were more likely to be single, employed in the formal sector and more highly educated than female household heads not cooking with PAYG LPG (Table 2). As previous research has found that more highly educated women are more likely to switch to clean cooking fuels (Andadari et al., 2014), additional research is needed to assess how PAYG LPG can reach the poorest households in informal settlements to prevent increased inequities in access to clean cooking fuels (Shupler et al., 2022b).

Conversely, a higher education level was not associated with uptake of PAYG LPG by women living in a male-run household (Table 2). This suggests that women that wish to transition to LPG in male-run households may sometimes lack the authority to switch their household’s primary cooking fuel. There is a growing body of research illustrating a positive association between women’s ability to participate in household decision-making and their likelihood of adopting LPG for cooking (Choudhuri and Desai, 2020; Gould and Urpelainen, 2020). Thus, while women’s education is key for increasing their likelihood of cooking with clean fuels (Kumar, 2018), other cultural norms may limit women’s ability to transition to clean energy (Kiptot et al., 2014).

### Use of time and financial savings

Time savings from switching to PAYG LPG for cooking was universally reported by women that previously cooked primarily with polluting fuels in the study setting. Regained time was due to both reductions in cooking and fuel collection time in the informal urban settlement (Figure 2). The average reduction in cooking time (42 min/day) reported by women previously cooking with polluting fuels is consistent with that of other studies (Krishnapriya et al., 2021; Simkovich et al., 2019).

Household dynamics appeared to further influence how women use their time and monetary savings. Women that were the household head were more likely to use their gained time to take on new employment; women that lived in households run by their male partner were more likely to fill their time completing household tasks (Figure 3). This further illustrates how women’s agency can influence their degree of economic gains as a result of having additional time to engage in other activites (Akter and Pratap, 2022; Nguyen and Su, 2021). We note that time savings were rarely used for educational purposes or leisure in this setting; a lack of evidence on the use of saved time for education has been documented by another study (Petrokofsky et al., 2021).

Despite female headed households having a lower average household income than male-run households (Table 1), a greater proportion of married female household heads were able to use their monetary savings from cooking with PAYG LPG for investment (41%) or savings (35%), compared with married women that were not household heads (3% and 21%, respectively) (Figure 4). Unmarried female household heads, whose household income was lower than that of married women, were still more likely than married women in male headed households to redirect their financial savings for investment (9%) or savings (23%). These findings have been echoed in an Indian study, which concluded that the positive externalities of LPG adoption are less likely to be realized in households where women do not participate in important household decision-making (Akter and Pratap, 2022).

As time savings have been identified as one of the main reasons that individuals adopt clean cooking fuels (Petrokofsky et al., 2021), increasing women’s awareness about the time savings and therefore increased productivity offered by cooking with clean fuels could increase their adoption faster than spreading knowledge about health or climate impacts. This is supported by the finding that the convenience of PAYG LPG was rated by nearly as many participants as a reason for recommending PAYG LPG to others (73%) as the ability to pay in smaller payments (83%); ‘better health’ was the least common reason (29%) stated for recommending PAYG LPG to others in the community.

### Added benefits of pay-as-you-go LPG over full cylinder LPG

Four-fifths (82%) of women previously cooking with full cylinder LPG reported saving time when switching to PAYG LPG. This is one of the first studies to document additional time savings (20 minutes/day) when women shift from full cylinder LPG to PAYG LPG. This 20 minute/day decline may be partially attributed to a reduction in the concurrent use of polluting fuels (fuel stacking) as PAYG LPG users were twice as likely to exclusively cook with LPG (61%) as those cooking with full cylinder LPG (33%) in the informal settlement (Table 2). This decline in stove stacking is plausibly due to PayGo Energy’s customers’ changing from single-burner to double-burner LPG stoves and thus having the ability to cook multiple meals simultaneously when using PAYG LPG. Somewhat unique to the Kenyan context is that the most common set up among full cylinder LPG users is a single-burner (‘meko’) stove (burner on top of the cylinder) due to its lower cost relative to double burner stoves (GLPGP, 2018; Shupler et al., 2022b).

In previous studies, many participants specifically referenced the switch from the single-burner ‘meko’ LPG stove to the double-burner LPG stove included as part of PayGo Energy’s package as the reason for faster cooking time and increased cost-effectiveness of PAYG LPG (Perros et al., 2021; Shupler et al., 2021c). Previous research conducted among multiple peri-urban communities in sub-Saharan Africa has further found that a higher number of LPG stoves burners is associated with increased per capita LPG consumption, regardless of socioeconomic status (Shupler et al., 2021a). Further, a study carried out in an informal settlement in Western Kenya highlighted that a greater number of stove burners per capita was associated with increased likelihood of sustaining use of LPG while under COVID-19 restrictions (Shupler et al., 2022b).

A finding unique to this urban setting was a 5-minute longer travel time required to obtain cooking fuels among households previously cooking with full cylinder LPG (33 min/trip) compared with those formerly cooking with polluting fuels (28 min/trip). The cylinder home deliveries offered by PayGo Energy therefore saved women that previously cooked with full cylinder LPG an average of 23 minutes/trip (Figure 2). Previous research conducted in an informal settlement in Western Kenyan identified that households using full cylinder LPG that received refills delivered to their home had a greater likelihood of using LPG as a primary fuel rather than a secondary fuel (Shupler et al., 2022b). The ability to have cylinders delivered directly to the home can also eliminate the risk of injury when collecting fuel, which has been associated with lower physical well-being in sub-Saharan Africa (Shupler et al., 2022a).

Moreover, financial savings were more common among women that switched from full cylinder LPG to PAYG LPG (59%) than among women previously cooking with polluting fuels (49%). Thus, despite the premium charged on LPG fuel under a PAYG LPG model, the ability to pay in smaller increments is economically beneficial to most women that formerly cooked with full cylinder LPG.

### Dietary changes

An improvement in dietary diversity and increased consumption of carbohydrates and protein-rich foods due to cooking with PAYG LPG was more prevalent among women-led households compared with households where males had authority over household decisions (Figure 5). Specifically, female primary cooks that were the household head were three times more likely to increase their household’s consumption of meat, fish and legumes compared with women that were not the head of household (Figure 5). While the mechanisms through which women’s agency may affect nutritional security are complex (Galiè et al., 2019), research has shown that women with greater control over household resources allows them greater flexibility to redirect their spending and consumption on food expenses, leading to improvements in diet (Doss, 2006; Duflo, 2003; Johnston et al., 2015). As the proportion of female household heads that used their time savings for grocery shopping (17%) was twice as high as that among women without household decision-making authority (8%), its plausible that women’s agency increased their ability to purchase more diverse and nutritious foods (Akter et al., 2017; Kadiyala et al., 2014; Sraboni et al., 2014). Multiple studies conducted in Ghana have found that women’s participation in household decision-making is associated with higher dietary diversity (Amugsi et al., 2016; Malapit and Quisumbing, 2015). Given that micronutrient deficiencies and undernutrition are part of the triple burden of malnutrition experienced by many families in sub-Saharan Africa (Hwalla et al., 2016; Swinburn et al., 2019), improved nutritional security as a result of cooking with modern energy can be an essential co-benefit of clean cooking access.

In addition to nutritional security, Women with control over food expenditures may also be more likely to increase the amount of food available to their families when cooking with PAYG LPG as households headed by women that were customers of PayGo Energy had a lower prevalence of food insecurity (54%) than female-run households in the general community (63%) (Table 2). This finding also can have significant health implications as households headed by women generally experience greater rates of food insecurity than households run by men (Kassie et al., 2015, 2014; Ragasa et al., 2019).

### Stress

Although a substantial proportion (70%) of women reported that their life was stressful (Table 3), rates of stress were approximately 10% lower among women cooking with PAYG LPG than in the general community (Table 3). Nearly two-thirds (64%) of female primary cooks using PAYG LPG reported that PAYG LPG reduced the amount of stress in their lives (Table 3).

This study builds upon other studies conducted in peri-urban sub-Saharan Africa (Shupler et al., 2022a) and rural India (Malakar and Day, 2020) that have found an association between cooking with clean fuels and mental health by elucidating potential pathways by which access to PAYG LPG for cooking may improve well-being. It was revealed that gaining additional time from cooking with clean fuels can empower women to take on new jobs to increase their income and spend more time buying food for their families (Bridge et al., 2016; Winter et al., 2019). Another study in China uncovered that increased participation in the labor force was a pathway by which access to clean cooking improved mental health (Liu et al., 2022). Thus, it is likely that increased productivity gained from clean cooking access can reduce stress associated with financial and food insecurity and therefore improve women’s well-being (O’Laughlin, 2007; Stevano, 2019).

Cooking with PAYG LPG was associated with lower rates of stress due to food and financial insecurity among female household heads, but not among women living in male headed households (Figure 6); for example, 52% of female household heads cooking with PAYG LPG reported that household expenses were a major source of stress, compared with 69% of female household heads not cooking with PAYG LPG. Conversely, among female primary cooks that were not the household head, the proportion reporting stress due to household expenses was larger among PAYG LPG users (55%) than those not cooking with PAYG LPG (46%) (Figure 6).

Due to their direct involvement in the management of household resources, females in charge of household decision making may have experienced greater reductions in stress as they may be in a better position to repurpose the additional money made available for household spending and consumption due to use of PAYG LPG (Richardson et al., 2019; Yount et al., 2014). Moreover, with female household heads being more inclined to prioritize their spending on food and clean cooking fuels, they may be more likely to sustain the benefits of PAYG LPG for a longer period of time; previous research among PayGo Energy’s customers found that female headed households were less likely to deactivate their PAYG LPG account prior to the COVID-19 pandemic than households with males in charge of cooking decisions (Shupler et al., 2021c).

Finally, almost half (45%) of women cooking with PAYG LPG reported that cooking with the technology allowed them to fulfill other aspirations which they could not pursue previously (Table 3). While the proportion of women able to fulfill previously unmet aspirations was higher among those that had never previously cooked with LPG (65%), the proportion of women previously cooking with full cylinder LPG that fulfilled previously unattainable aspirations was considerably high (38%).

### Strengths and limitations

This study is one of the first to document how women use their time savings when cooking with PAYG LPG. Differences in cooking patterns between PayGo Energy customers and the general community, such as a lower rate of fuel stacking among those using PAYG LPG versus full cylinder LPG (Table 2), may be attributed to varying levels of income and not entirely to the PAYG model itself. Longitudinal studies are needed to elucidate potential reductions in polluting cooking fuel use when gaining access to PAYG LPG.

As this study was conducted in a community with a relatively high rate of existing use of full cylinder LPG (three-quarters of PayGo Energy’s customers previously cooked with full cylinder LPG), the use of PAYG LPG and perceived benefits may differ in a community with a much lower penetration of full cylinder LPG. Similarly, the results from this informal urban settlement may not be generalizable to other settings, including rural communities; PAYG LPG has not expanded to rural areas in sub-Saharan Africa due to increased distribution challenges and transportation costs. While this study was conducted in a single urban area, the connection between access to clean energy, women’s empowerment and improved well-being has been founded across many contexts (Fejerskov, 2017; Sehjpal et al., 2014).

It is important to carry out similar studies in rural areas where time poverty due to procuring fuelwood may represent a significantly higher time sink for women. It is possible that the gendered differences in time savings detected in this study may therefore be conservative relative to rural locations. Nonetheless, this study confirms that PAYG LPG can save women time even in urban areas where fuel collection times are typically no more than 30 minutes/trip.

As this study and previous research among PayGo Energy’s customers (Shupler et al., 2021c) has shown that women cooking with PAYG LPG are generally of higher SES than the overall community, it is possible that this study sample of are not representative of Nairobi’s informal settlement population. Thus, women’s socioeconomic benefits reported in this study may slightly differ from women of different socioeconomic positions. Even so, the universal time savings reported by women suggests that social benefits will likely manifest for women across all income brackets and education levels.

There is the potential for social desirability bias to exist in the survey responses as participants knew they were being questioned about PAYG LPG. Additionally, the comparative sample of primary cooks that were not customers of PayGo Energy was relatively smaller than those using PAYG LPG, which may have led to lower reliability. However, we note that the household income and education levels of the random sample is similar to that of a study conducted among a larger sample of 474 primary cooks in the same informal settlement (Shupler et al., 2021b).

As information on stress levels and diet quality was self-reported in this study, there is potential for bias. Nonetheless, the results are likely robust given the substantial literature documenting associations between food and nutritional security with reductions in stress (O’Laughlin, 2007; Stevano, 2019).

## CONCLUSION

This study highlights the important role that women’s agency has on the social and economic benefits they achieve when transitioning to PAYG LPG for cooking. Compared to women with less authority in household decision-making, female household heads were more likely to experience improvements in diet, reductions in stress levels, greater employment opportunities and a greater ability to save or invest their financial savings from being a PayGo Energy customer.

### Policy considerations

Given that women’s power within the household plays an important role in their realized benefits from adoption of PAYG LPG, household power dynamics, including how responsibilities for household tasks change with transitions to clean fuels, should increasingly be examined in future research and factored into clean cooking interventions, communication campaigns and implementation approaches. This will allow for better tracking of the expected impact of clean household energy policies on gender equality (SDG target 5c) and illustrate women’s progress toward accessing household energy resources to support their empowerment (SDG target 5b) (Haddad et al., 2021). A better understanding of how women’s position influences their perceived socioeconomic benefits when gaining access to LPG will also help increase the likelihood of sustained use of clean cooking fuels (Broto et al., 2017).

Moreover, this study uncovers measurable benefits of adoption of PAYG LPG that are above that of full cylinder LPG. These benefits, such as further time savings, are not attributed solely to the ability to pay in small increments, but also the convenience of cylinder home deliveries and access to double burner stoves. As socioeconomic benefits are likely to have an immediate, visible impact on women’s well-being compared with reductions in household air pollution exposure and emissions of climate-forcing pollutants, it will likely be beneficial for clean fuel programs and interventions to be marketed for their timesaving capabilities (Petrokofsky et al., 2021; Troncoso et al., 2019).

Finally, given the positive impacts that access to PAYG LPG has on women’s livelihoods, there is a need to expand data collection on the gendered benefits of clean cooking fuel adoption. This includes developing novel, quantifiable measures that relate clean cooking to gender equality, such as ‘quality hours’ (additional time spent on productive activities, education or leisure) gained by women (Modern Energy Cooking Services and Energy 4 Impact, 2022). These metrics can be used to support innovative, results-based funding mechanisms for such as Development Impact Bonds (e.g. ‘SDG 5 Impact Statements’ (The Gold Standard, 2022)) that will attract much-needed investment into the clean cooking sector and advance gender equality.

## Data Availability

All data produced in the present study are available upon reasonable request to the authors.

## Acknowledgments

The authors would like to sincerely express gratitude to the field team for their contribution to study implementation. This study was funded by the Shell Foundation. Matthew Shupler is funded by Wellcome Trust and National Institute for Health Research.

## Competing Interests

Mark O’Keefe is co-founder and Product Manager at *PayGo Energy*. His employment at *PayGo Energy* had no impact on interpretation of the data.

## Notes

### Competing Interest Statement

Mark OKeefe is co-founder and Product Manager at PayGo Energy. His employment at PayGo Energy had no impact on interpretation of the data.

### Author Declarations

Ethics committee of National Commission for Science, Technology & Innovation gave ethical approval for this work

## References

Aboderin, I., Kano, M., Owii, H.A., 2017. Toward “Age-Friendly Slums”? Health Challenges of Older Slum Dwellers in Nairobi and the Applicability of the Age-Friendly City Approach. International Journal of Environmental Research and Public Health 14, 1259. https://doi.org/10.3390/ijerph14101259

Ahmed, S., International Institute for Environment and Development, Human Settlements Programme, 2015. Cooking up a storm: community-led mapping and and advocacy with food vendors in Nairobi’s informal settlements.

Akter, S., Pratap, C., 2022. Impact of clean cooking fuel adoption on women’s welfare in India: the mediating role of women’s autonomy. Sustain Sci 17, 243–257. https://doi.org/10.1007/s11625-021-01069-9

Akter, S., Rutsaert, P., Luis, J., Htwe, N.M., San, S.S., Raharjo, B., Pustika, A., 2017. Women’s empowerment and gender equity in agriculture: A different perspective from Southeast Asia. Food Policy 69, 270–279. https://doi.org/10.1016/j.foodpol.2017.05.003

Alexander, D., Northcross, A., Wilson, N., Dutta, A., Pandya, R., Ibigbami, T., Adu, D., Olamijulo, J., Morhason-Bello, O., Karrison, T., Ojengbede, O., Olopade, C.O., 2017. Randomized Controlled Ethanol Cookstove Intervention and Blood Pressure in Pregnant Nigerian Women. American Journal of Respiratory and Critical Care Medicine 195, 1629–1639. https://doi.org/10.1164/rccm.201606-1177OC

Alexander, D.A., Northcross, A., Karrison, T., Morhasson-Bello, O., Wilson, N., Atalabi, O.M., Dutta, A., Adu, D., Ibigbami, T., Olamijulo, J., Adepoju, D., Ojengbede, O., Olopade, C.O., 2018. Pregnancy outcomes and ethanol cook stove intervention: A randomized-controlled trial in Ibadan, Nigeria. Environment International 111, 152–163. https://doi.org/10.1016/j.envint.2017.11.021

Amugsi, D.A., Lartey, A., Kimani-Murage, E., Mberu, B.U., 2016. Women’s participation in household decision-making and higher dietary diversity: findings from nationally representative data from Ghana. Journal of Health, Population and Nutrition 35, 16. https://doi.org/10.1186/s41043-016-0053-1

Andadari, R.K., Mulder, P., Rietveld, P., 2014. Energy poverty reduction by fuel switching. Impact evaluation of the LPG conversion program in Indonesia. Energy Policy 66, 436–449. https://doi.org/10.1016/j.enpol.2013.11.021

Anenberg, S.C., Balakrishnan, K., Jetter, J., Masera, O., Mehta, S., Moss, J., Ramanathan, V., 2013. Cleaner Cooking Solutions to Achieve Health, Climate, and Economic Cobenefits. Environ. Sci. Technol. 47, 3944–3952. https://doi.org/10.1021/es304942e

Arku, R.E., Ezzati, M., Baumgartner, J., Fink, G., Zhou, B., Hystad, P., Brauer, M., 2018. Elevated blood pressure and household solid fuel use in premenopausal women: Analysis of 12 Demographic and Health Surveys (DHS) from 10 countries. Environmental Research 160, 499–505. https://doi.org/10.1016/j.envres.2017.10.026

Bailis, R., Drigo, R., Ghilardi, A., Masera, O., 2015. The carbon footprint of traditional woodfuels. Nature Clim Change 5, 266–272. https://doi.org/10.1038/nclimate2491

Bates, M.N., Chandyo, R.K., Valentiner-Branth, P., Pokhrel, A.K., Mathisen, M., Basnet, S., Shrestha, P.S., Strand, T.A., Smith, K.R., 2013. Acute Lower Respiratory Infection in Childhood and Household Fuel Use in Bhaktapur, Nepal. Environmental Health Perspectives 121, 637–642. https://doi.org/10.1289/ehp.1205491

Beguy, D., Elung’ata, P., Mberu, B., Oduor, C., Wamukoya, M., Nganyi, B., Ezeh, A., 2015. Health & Demographic Surveillance System Profile: The Nairobi Urban Health and Demographic Surveillance System (NUHDSS). International Journal of Epidemiology 44, 462–471. https://doi.org/10.1093/ije/dyu251

Bridge, B.A., Adhikari, D., Fontenla, M., 2016. Electricity, income, and quality of life. The Social Science Journal 53, 33–39. https://doi.org/10.1016/j.soscij.2014.12.009

Broto, V.C., Stevens, L., Ackom, E., Tomei, J., Parikh, P., Bisaga, I., To, L.S., Kirshner, J., Mulugetta, Y., 2017. A research agenda for a people-centred approach to energy access in the urbanizing global south. Nat Energy 2, 776–779. https://doi.org/10.1038/s41560-017-0007-x

Bruce, N., Aunan, K., Rehfuess, E., 2017. Liquefied petroleum gas as a clean cooking fuel for developing countries: implications for climate, forests, and affordability. KfW Development Bank.

Bruce, N., de Cuevas, R.A., Cooper, J., Enonchong, B., Ronzi, S., Puzzolo, E., MBatchou, B., Pope, D., 2018. The Government-led initiative for LPG scale-up in Cameroon: Programme development and initial evaluation. Energy for Sustainable Development 46, 103–110. https://doi.org/10.1016/j.esd.2018.05.010

Burroughs Pena, M., Romero, K.M., Velazquez, E.J., Davila-Roman, V.G., Gilman, R.H., Wise, R.A., Miranda, J.J., Checkley, W., 2015. Relationship Between Daily Exposure to Biomass Fuel Smoke and Blood Pressure in High-Altitude Peru. Hypertension 65, 1134–1140. https://doi.org/10.1161/HYPERTENSIONAHA.114.04840

Choudhuri, P., Desai, S., 2020. Gender inequalities and household fuel choice in India. Journal of Cleaner Production 265, 121487. https://doi.org/10.1016/j.jclepro.2020.121487

Clean Cooking Alliance, 2019. Scaling LPG For Cooking in Developing Markets: Insights from Tanzania. Washington (DC).

Dianati, K., Zimmermann, N., Milner, J., Muindi, K., Ezeh, A., Chege, M., Mberu, B., Kyobutungi, C., Fletcher, H., Wilkinson, P., Davies, M., 2019. Household air pollution in Nairobi’s slums: A long-term policy evaluation using participatory system dynamics. Science of The Total Environment 660, 1108–1134. https://doi.org/10.1016/j.scitotenv.2018.12.430

Doss, C., 2006. The Effects of Intrahousehold Property Ownership on Expenditure Patterns in Ghana. Journal of African Economies 15, 149–180. https://doi.org/10.1093/jae/eji025

Duflo, E., 2003. Grandmothers and Granddaughtersl: Old-Age Pensions and Intrahousehold Allocation in South Africa. World Bank Economic Review 17, 1–25. https://doi.org/10.1093/wber/lhg013

Fatmi, Z., Ntani, G., Coggon, D., 2019. Coronary heart disease, hypertension and use of biomass fuel among women: comparative cross-sectional study. BMJ Open 9, e030881. https://doi.org/10.1136/bmjopen-2019-030881

Fejerskov, A.M., 2017. The Influence of Established Ideas in Emerging Development Organisations: Gender Equality and the Bill and Melinda Gates Foundation. The Journal of Development Studies 53, 584–599. https://doi.org/10.1080/00220388.2016.1199859

French Gates, M., 2014. Putting women and girls at the center of development. Science 345, 1273–1275. https://doi.org/10.1126/science.1258882

Galiè, A., Teufel, N., Girard, A.W., Baltenweck, I., Dominguez-Salas, P., Price, M.J., Jones, R., Lukuyu, B., Korir, L., Raskind, IlanaG., Smith, K., Yount, K.M., 2019. Women’s empowerment, food security and nutrition of pastoral communities in Tanzania. Global Food Security 23, 125–134. https://doi.org/10.1016/j.gfs.2019.04.005

Gould, C.F., Urpelainen, J., 2020. The Gendered Nature of Liquefied Petroleum Gas Stove Adoption and Use in Rural India. The Journal of Development Studies 56, 1309–1329. https://doi.org/10.1080/00220388.2019.1657571

Haddad, Z., Williams, K.N., Lewis, J.J., Prats, E.V., Adair-Rohani, H., 2021. Expanding data is critical to assessing gendered impacts of household energy use. BMJ 375, n2273. https://doi.org/10.1136/bmj.n2273

Haushofer, J., Collins, M., de Giusti, G., Njoroge, J., Odero, A., Onyango, C., Vancel, J., Jang, C., Kuruvilla, M., Hughes, C., 2014. A Methodology for Laboratory Experiments in Developing Countries: Examples from the Busara Center (SSRN Scholarly Paper No. 2155217). Social Science Research Network, Rochester, NY. https://doi.org/10.2139/ssrn.2155217

Hosonuma, N., Herold, M., De Sy, V., De Fries, R.S., Brockhaus, M., Verchot, L., Angelsen, A., Romijn, E., 2012. An assessment of deforestation and forest degradation drivers in developing countries. Environ. Res. Lett. 7, 044009. https://doi.org/10.1088/1748-9326/7/4/044009

Jagoe, K., Rossanese, M., Charron, D., Rouse, J., Waweru, F., Waruguru, M., Delapena, S., Piedrahita, R., Livingston, K., Ipe, J., 2020. Sharing the burden: Shifts in family time use, agency and gender dynamics after introduction of new cookstoves in rural Kenya. Energy Research & Social Science 64, 101413. https://doi.org/10.1016/j.erss.2019.101413

Johnson, O.W., Gerber, V., Muhoza, C., 2019. Gender, culture and energy transitions in rural Africa. Energy Research & Social Science 49, 169–179. https://doi.org/10.1016/j.erss.2018.11.004

Johnston, D., Stevano, S., Malapit, H.J., Hull, E., Kadiyala, S., 2015. Agriculture, Gendered Time Use, and Nutritional Outcomes: A Systematic Review (SSRN Scholarly Paper No. 2685291). Social Science Research Network, Rochester, NY.

Kadiyala, S., Harris, J., Headey, D., Yosef, S., Gillespie, S., 2014. Agriculture and nutrition in India: mapping evidence to pathways. Ann N Y Acad Sci 1331, 43–56. https://doi.org/10.1111/nyas.12477

Kassie, M., Ndiritu, S.W., Stage, J., 2014. What Determines Gender Inequality in Household Food Security in Kenya? Application of Exogenous Switching Treatment Regression. World Development 56, 153–171. https://doi.org/10.1016/j.worlddev.2013.10.025

Kassie, M., Stage, J., Teklewold, H., Erenstein, O., 2015. Gendered food security in rural Malawi: why is women’s food security status lower? Food Sec. 7, 1299–1320. https://doi.org/10.1007/s12571-015-0517-y

Kim, H.-S., Yoon, Y., Mutinda, M., 2019. Secure land tenure for urban slum-dwellers: A conjoint experiment in Kenya. Habitat International 93, 102048. https://doi.org/10.1016/j.habitatint.2019.102048

Kiptot, E., Franzel, S., Degrande, A., 2014. Gender, agroforestry and food security in Africa. Current Opinion in Environmental Sustainability, Sustainability challenges 6, 104–109. https://doi.org/10.1016/j.cosust.2013.10.019

Krishnapriya, P.P., Chandrasekaran, M., Jeuland, M., Pattanayak, S.K., 2021. Do improved cookstoves save time and improve gender outcomes? Evidence from six developing countries. Energy Economics 102, 105456. https://doi.org/10.1016/j.eneco.2021.105456

Kumar, A., 2018. Justice and politics in energy access for education, livelihoods and health: How socio-cultural processes mediate the winners and losers. Energy Research & Social Science 40, 3–13. https://doi.org/10.1016/j.erss.2017.11.029

Kumar, P., Mehta, S., 2016. Poverty, gender, and empowerment in sustained adoption of cleaner cooking systems: Making the case for refined measurement. Energy Research & Social Science 19, 48–52. https://doi.org/10.1016/j.erss.2016.05.018

Kurmi, O.P., Devereux, G.S., Smith, W.C.S., Semple, S., Steiner, M.F.C., Simkhada, P., Lam, K.-B.H., Ayres, J.G., 2013. Reduced lung function due to biomass smoke exposure in young adults in rural Nepal. European Respiratory Journal 41, 25–30. https://doi.org/10.1183/09031936.00220511

Kypridemos, C., Puzzolo, E., Aamaas, B., Hyseni, L., Shupler, M., Aunan, K., Pope, D., 2020. Health and Climate Impacts of Scaling Adoption of Liquefied Petroleum Gas (LPG) for Clean Household Cooking in Cameroon: A Modeling Study. Environ Health Perspect 128, 047001. https://doi.org/10.1289/EHP4899

Maji, P., Mehrabi, Z., Kandlikar, M., 2021. Incomplete transitions to clean household energy reinforce gender inequality by lowering women’s respiratory health and household labour productivity. World Development 139, 105309. https://doi.org/10.1016/j.worlddev.2020.105309

Malakar, Y., Day, R., 2020. Differences in firewood users’ and LPG users’ perceived relationships between cooking fuels and women’s multidimensional well-being in rural India. Nat Energy 5, 1022–1031. https://doi.org/10.1038/s41560-020-00722-4

Malapit, H.J.L., Kadiyala, S., Quisumbing, A.R., Cunningham, K., Tyagi, P., 2015. Women’s Empowerment Mitigates the Negative Effects of Low Production Diversity on Maternal and Child Nutrition in Nepal. The Journal of Development Studies 51, 1097–1123. https://doi.org/10.1080/00220388.2015.1018904

Malapit, H.J.L., Quisumbing, A.R., 2015. What dimensions of women’s empowerment in agriculture matter for nutrition in Ghana? Food Policy 52, 54–63. https://doi.org/10.1016/j.foodpol.2015.02.003

Mazorra, J., Sánchez-Jacob, E., de la Sota, C., Fernández, L., Lumbreras, J., 2020. A comprehensive analysis of cooking solutions co-benefits at household level: Healthy lives and well-being, gender and climate change. Science of The Total Environment 707, 135968. https://doi.org/10.1016/j.scitotenv.2019.135968

Mberu, B.U., Haregu, T.N., Kyobutungi, C., Ezeh, A.C., 2016. Health and health-related indicators in slum, rural, and urban communities: a comparative analysis. Glob Health Action 9. https://doi.org/10.3402/gha.v9.33163

Musango, J.K., 2022. Assessing gender and energy in urban household energy transitions in South Africa: A quantitative storytelling from Groenheuwel informal settlement. Energy Research & Social Science 88, 102525. https://doi.org/10.1016/j.erss.2022.102525

Musango, J.K., Smit, S., Ceschin, F., Ambole, A., Batinge, B., Anditi, C., Petrulaityte, A., Mukama, M., 2020. Mainstreaming gender to achieve security of energy services in poor urban environments. Energy Research & Social Science 70, 101715. https://doi.org/10.1016/j.erss.2020.101715

Nguyen, C.P., Su, T.D., 2021. Does energy poverty matter for gender inequality? Global evidence. Energy for Sustainable Development 64, 35–45. https://doi.org/10.1016/j.esd.2021.07.003

O’Keefe, M., Marcigot, F., 2020. IoT LPG Cylinder Tag & Trace, MECS-TRIID Project Report. Modern Energy Cooking Services (MECS).

O’Laughlin, B., 2007. A Bigger Piece of a Very Small Pie: Intrahousehold Resource Allocation and Poverty Reduction in Africa. Development and Change 38, 21–44. https://doi.org/10.1111/j.1467-7660.2007.00401.x

Parikh, J., 2011. Hardships and health impacts on women due to traditional cooking fuels: A case study of Himachal Pradesh, India. Energy Policy, Clean Cooking Fuels and Technologies in Developing Economies 39, 7587–7594. https://doi.org/10.1016/j.enpol.2011.05.055

Parikh, P., Chaturvedi, S., George, G., 2012. Empowering change: The effects of energy provision on individual aspirations in slum communities. Energy Policy, Special Section: Past and Prospective Energy Transitions - Insights from History 50, 477–485. https://doi.org/10.1016/j.enpol.2012.07.046

Parikh, P., Fu, K., Parikh, H., McRobie, A., George, G., 2015. Infrastructure Provision, Gender, and Poverty in Indian Slums. World Development 66, 468–486. https://doi.org/10.1016/j.worlddev.2014.09.014

Paul, L.A., Burnett, R.T., Kwong, J.C., Hystad, P., van Donkelaar, A., Bai, L., Goldberg, M.S., Lavigne, E., Copes, R., Martin, R.V., Kopp, A., Chen, H., 2020. The impact of air pollution on the incidence of diabetes and survival among prevalent diabetes cases. Environ Int 134, 105333. https://doi.org/10.1016/j.envint.2019.105333

Perros, T., Büttner, P., Leary, J., Parikh, P., 2021. Pay-as-you-go LPG: A mixed-methods pilot study in urban Rwanda. Energy for Sustainable Development 65, 117–129. https://doi.org/10.1016/j.esd.2021.10.003

Petrokofsky, G., Harvey, W.J., Petrokofsky, L., Ochieng, C.A., 2021. The Importance of Time-Saving as a Factor in Transitioning from Woodfuel to Modern Cooking Energy Services: A Systematic Map. Forests 12, 1149. https://doi.org/10.3390/f12091149

Prabhu, V.S., 2010. Tests of Intrahousehold Resource Allocation Using a CV Framework: A Comparison of Husbands’ and Wives’ Separate and Joint WTP in the Slums of Navi-Mumbai, India. World Development 38, 606–619. https://doi.org/10.1016/j.worlddev.2009.12.002

R Core Team, 2017. R: A language and environment for statistical computing. R Foundation for Statistical Computing, Vienna, Austria.

Ragasa, C., Aberman, N.-L., Alvarez Mingote, C., 2019. Does providing agricultural and nutrition information to both men and women improve household food security? Evidence from Malawi. Global Food Security 20, 45–59. https://doi.org/10.1016/j.gfs.2018.12.007

Richardson, R.A., Harper, S., Bates, L.M., Nandi, A., 2019. The effect of agency on women’s mental distress: A prospective cohort study from rural Rajasthan, India. Social Science & Medicine 233, 47–56. https://doi.org/10.1016/j.socscimed.2019.05.052

Sehjpal, R., Ramji, A., Soni, A., Kumar, A., 2014. Going beyond incomes: Dimensions of cooking energy transitions in rural India. Energy 68, 470–477. https://doi.org/10.1016/j.energy.2014.01.071

Shupler, M., Baame, M., Nix, E., Tawiah, T., Lorenzetti, F., Saah, J., Anderson de Cuevas, R., Sang, E., Puzzolo, E., Mangeni, J., Betang, E., Twumasi, M., Amenga-Etego, S., Quansah, R., Mbatchou, B., Menya, D., Asante, K.P., Pope, D., 2022a. Multiple aspects of energy poverty are associated with lower mental health-related quality of life: A modelling study in three peri-urban African communities. SSM - Mental Health 2, 100103. https://doi.org/10.1016/j.ssmmh.2022.100103

Shupler, M., Mangeni, J., Tawiah, T., Sang, E., Baame, M., Anderson de Cuevas, R., Nix, E., Betang, E., Saah, J., Twumasi, M., Amenga-Etego, S., Quansah, R., Puzzolo, E., Mbatchou, B., Asante, K.P., Menya, D., Pope, D., 2021a. Modelling of supply and demand-side determinants of liquefied petroleum gas consumption in peri-urban Cameroon, Ghana and Kenya. Nat Energy 1–13. https://doi.org/10.1038/s41560-021-00933-3

Shupler, M., Menya, D., Sang, E., Anderson de Cuevas, R., Mang’eni, J., Lorenzetti, F., Saligari, S., Nix, E., Mwitari, J., Gohole, A., Pope, D., Puzzolo, E., 2022b. Widening inequities in clean cooking fuel use and food security: compounding effects of COVID-19 restrictions & VAT on LPG in a Kenyan informal urban settlement. Environ. Res. Lett. https://doi.org/10.1088/1748-9326/ac6761

Shupler, M., Mwitari, J., Gohole, A., Anderson de Cuevas, R., Puzzolo, E., Cukic, I., Nix, E., Pope, D., 2021b. COVID-19 impacts on household energy & food security in a Kenyan informal settlement: The need for integrated approaches to the SDGs. Renewable and Sustainable Energy Reviews 144, 111018. https://doi.org/10.1016/j.rser.2021.111018

Shupler, M., O’Keefe, M., Puzzolo, E., Nix, E., Anderson de Cuevas, R., Mwitari, J., Gohole, A., Sang, E., Cukic, I., Menya, D., Pope, D., 2021c. Pay-as-you-go liquefied petroleum gas supports sustainable clean cooking in Kenyan informal urban settlement during COVID-19 lockdown. Applied Energy 292, 116769. https://doi.org/10.1016/j.apenergy.2021.116769

Siddharthan, T., Grigsby, M.R., Goodman, D., Chowdhury, M., Rubinstein, A., Irazola, V., Gutierrez, L., Miranda, J.J., Bernabe-Ortiz, A., Alam, D., Kirenga, B., Jones, R., van Gemert, F., Wise, R.A., Checkley, W., 2018. Association between Household Air Pollution Exposure and Chronic Obstructive Pulmonary Disease Outcomes in 13 Low- and Middle-Income Country Settings. Am. J. Respir. Crit. Care Med. 197, 611–620. https://doi.org/10.1164/rccm.201709-1861OC

Simkovich, S.M., Williams, K.N., Pollard, S., Dowdy, D., Sinharoy, S., Clasen, T.F., Puzzolo, E., Checkley, W., 2019. A Systematic Review to Evaluate the Association between Clean Cooking Technologies and Time Use in Low- and Middle-Income Countries. International Journal of Environmental Research and Public Health 16, 2277. https://doi.org/10.3390/ijerph16132277

Singh, D., Pachauri, S., Zerriffi, H., 2017. Environmental payoffs of LPG cooking in India. Environ. Res. Lett. 12, 115003. https://doi.org/10.1088/1748-9326/aa909d

Smith, K.R., McCracken, J.P., Weber, M.W., Hubbard, A., Jenny, A., Thompson, L.M., Balmes, J., Diaz, A., Arana, B., Bruce, N., 2011. Effect of reduction in household air pollution on childhood pneumonia in Guatemala (RESPIRE): a randomised controlled trial. The Lancet 378, 1717–1726. https://doi.org/10.1016/S0140-6736(11)60921-5

Sraboni, E., Malapit, H.J., Quisumbing, A.R., Ahmed, A.U., 2014. Women’s Empowerment in Agriculture: What Role for Food Security in Bangladesh? World Development 61, 11–52. https://doi.org/10.1016/j.worlddev.2014.03.025

Stevano, S., 2019. The Limits of Instrumentalism: Informal Work and Gendered Cycles of Food Insecurity in Mozambique. The Journal of Development Studies 55, 83–98. https://doi.org/10.1080/00220388.2017.1408793

Stoner, O., Lewis, J., Martínez, I.L., Gumy, S., Economou, T., Adair-Rohani, H., 2021. Household cooking fuel estimates at global and country level for 1990 to 2030. Nat Commun 12, 5793. https://doi.org/10.1038/s41467-021-26036-x

Thompson, L.M., Bruce, N., Eskenazi, B., Diaz, A., Pope, D., Smith, K.R., 2011. Impact of Reduced Maternal Exposures to Wood Smoke from an Introduced Chimney Stove on Newborn Birth Weight in Rural Guatemala. Environmental Health Perspectives 119, 1489–1494. https://doi.org/10.1289/ehp.1002928

Troncoso, K., Segurado, P., Aguilar, M., Soares da Silva, A., 2019. Adoption of LPG for cooking in two rural communities of Chiapas, Mexico. Energy Policy 133, 110925. https://doi.org/10.1016/j.enpol.2019.110925

UN-Habitat, 2013. Housing and Slum Upgrading; Gender issue guide.

Van Leeuwen, V., Evans, A., Hyseni, B., 2017. Increasing the use of liquefied petroleum gas in cooking in developing countries (No. 114846). The World Bank, LiveWire.

Winter, S.C., Obara, L.M., Barchi, F., 2019. Environmental Correlates of Health-Related Quality of Life among Women Living in Informal Settlements in Kenya. International Journal of Environmental Research and Public Health 16, 3948. https://doi.org/10.3390/ijerph16203948

Yasmin, N., Grundmann, P., 2020. Home-cooked energy transitions: Women empowerment and biogas-based cooking technology in Pakistan. Energy Policy 137, 111074. https://doi.org/10.1016/j.enpol.2019.111074

Yount, K.M., Dijkerman, S., Zureick-Brown, S., VanderEnde, K.E., 2014. Women’s empowerment and generalized anxiety in Minya, Egypt. Social Science & Medicine 106, 185–193. https://doi.org/10.1016/j.socscimed.2014.01.022

Yu, K., Qiu, G., Chan, K.-H., Lam, K.-B.H., Kurmi, O.P., Bennett, D.A., Yu, C., Pan, A., Lv, J., Guo, Y., Bian, Z., Yang, L., Chen, Y., Hu, F.B., Chen, Z., Li, L., Wu, T., 2018. Association of Solid Fuel Use With Risk of Cardiovascular and All-Cause Mortality in Rural China. JAMA 319, 1351–1361. https://doi.org/10.1001/jama.2018.2151

